# Cultural engagement is related to decelerated physiological age: doubly-robust estimations in a national cohort study

**DOI:** 10.1101/2025.10.23.25338631

**Authors:** Daisy Fancourt, Saoirse Finn, Hei Wan Mak, Andrew Steptoe, Mikaela Bloomberg

**Affiliations:** Department of Behavioural Science and Health, University College London; Department of Epidemiology and Public Health, University College London

## Abstract

Cultural engagement (e.g. going to live music events and theatre performances, museums, galleries and exhibitions, and the cinema) is longitudinally associated in repeated epidemiological studies with age-related mental and physical health outcomes. However, it is unclear whether it also influences how fast older adults age physiologically – so-called age acceleration. This study aimed to ascertain whether regular cultural engagement amongst older adults is related to slower physiological ageing using a previously-derived physiological age index and a doubly-robust estimation approach to account for confounders. Using older adults from the English Longitudinal Study of Ageing (n=4,467), we found that cultural engagement was related to lower physiological age cross-sectionally (average treatment effect −2.17; 95% CI −3.48 to −0.86), and 4 and 8 years later. The effect was seen consistently for all three types of cultural activity explored (cultural performances, museums/exhibitions, and the cinema). These analyses were robust to multiple sensitivity analyses, including considering alternative confounding structures, outliers, and treatment specification. Overall, these findings provide insight into how cultural engagement may be related to processes of ageing.

## Introduction

The arts encompass a diverse range of human practices relating to the production or experience of human creativity and imagination, including actively participating in the arts (e.g. dance, music, crafts; hereafter “arts participation”) and attending cultural events (e.g. going to live music events, museums, galleries, theatre performances, and the cinema; hereafter “cultural engagement”). Over the past two decades, increasing experimental and epidemiological research has identified diverse mental and physical health benefits that occur as a result of arts participation and cultural engagement, leading to calls for the arts to be formally recognised as a health behaviour.^1–3^ Theoretically, such effects may occur due to the ‘active ingredients’ arts participation and cultural engagement comprise and the mechanisms of action these ingredients activate. From an ingredients perspective, arts and cultural engagement act as a vehicle to well-known salutogenic behaviours such as social interaction, physical activity and cognitive stimulation. But they also bring people into contact with other ingredients such as creativity, imagination, multi-sensory stimulation and aesthetics that are strongly expressed in the arts.^4^ These ingredients synergistically activate diverse psychological, biological and behavioural mechanisms that are linked to health-related outcomes.^5^

Focusing on cultural engagement specifically, amongst older adults this behaviour has been longitudinally associated in repeated epidemiological studies with subsequent improvements in mental health and wellbeing, as well as reduced risk of developing age-related disability or impairment, chronic pain, frailty, chronic diseases like coronary heart disease, cognitive decline and dementia, and even premature mortality.^6–17^ These results have been found independent of measured and unmeasured confounders, using diverse statistical approaches that each account for different types of bias, with some of these findings corroborated by experimental evidence.^18,19^ However, while such evidence suggests that cultural engagement may positively influence age-related outcomes, what is less clear is whether it also influences how fast older adults age physiologically – so-called age acceleration.

Over the past decade, there has been increasing interest in the use of biological ageing clocks that combine age-related molecular features and quantify discrepancies between chronological vs biological age, enabling the calculation of whether individuals are experiencing accelerated or decelerated ageing. Some ageing clocks focus on DNA methylation data, while other clocks have been generated from downstream molecular phenotypes such as proteomic data and metabolomic data, or from broader clinical data.^20^ Physiological clocks are an example of a downstream biological clock that combine phenotypic measures of blood-based biomarkers (e.g. fibrinogen, C-reactive protein and glycated haemoglobin) with tests of physiological function (e.g. grip strength, respiratory function, pulse pressure and blood pressure) that have been found to be valuable in predicting age-related pathology.^21,22^ These types of clocks are of particular value as they are sometimes more strongly related to disease outcomes of interest than methylation-based clocks because they look at downstream impacts of cellular ageing more proximate to disease outcomes.^23^ In previous work, we have generated and validated one such physiological clock and demonstrated that physiological age relative to chronological age predicts many of the same age-related outcomes that cultural engagement has been related to, including cardiovascular disease, aspects of frailty, cognitive dysfunction, dementia and mortality.^24^ Significantly, in previous work relating deficits in social connections to this physiological clock, the strongest relationship with decelerated physiological age was for “social integration”, which comprised behaviours such as community group membership, volunteering and cultural engagement.^25^ However, as social integration is a broad index, it remains unclear whether cultural engagement could be one of the active drivers of the observed associations. This warrants further investigation given the benefits cultural engagement has been reported to have on mental and physical health in older adults.

Consequently, this study aimed to ascertain whether regular cultural engagement amongst older adults is related to slower physiological ageing. Our primary analyses (RQ1) explored associations between an index of cultural engagement and age acceleration/deceleration (calculated as the gap between an individual’s physiological age and their chronological age) both cross-sectionally and 4 and 8 years later. Our secondary analyses (RQ2) explored which type of cultural activity might be driving any potential associations. To improve causal inference, we designated a clear “treatment” group (i.e. those who engage regularly with cultural activities) and “control” group (i.e. those who never engage) and applied a doubly-robust estimation approach to account for the non-random treatment assignment.

## Methods

### Dataset

Data were derived from the English Longitudinal Study of Ageing (ELSA). ELSA is a large-scale panel study of people aged 50 and over and their partners, living in private households in England. The original sample was drawn from participants from the Health Survey in England (HSE) in 1998, 1999 and 2001.^26^ The first wave of data collection commenced in 2002/2003, and participants have been followed biennially since. We used wave 2 (2004/5), as this wave included a rich battery of cultural and physiological variables of relevance, with longitudinal follow-up data from nurse visits at wave 4 (2008/9) and wave 6 (2012/13). ELSA receives ethical approval from the NHS Research Ethics Service.

We restricted participants to core ELSA members who had returned the self-completion questionnaire where most of our main variables of interest were measured and were over the age of 50 at the point of questionnaire completion (n=8,129), who had physiological data available (from blood samples and nurse visits) to generate a physiological ageing index at wave 2 (n=5,572), and who had full data on exposures and confounders (n=5,349). Within this sample, we followed a potential outcomes framework approach and defined our exposure (cultural engagement) as a hypothetical intervention.^27,28^ So we designated our “treatment” group to be “attending one or more of the three cultural exposures every few months or more” (described further below; n=2,480) and a “control” group to be “never engaging” (n=870). We excluded participants who fell between these two thresholds of engagement (i.e. “less than once a year”) in order to emulate the conditions of a randomised trial more closely. This provided a total analytic sample size of n=3,350 (see Appendix 2 Supplementary Table 1 for sample size across waves).

**Table 1.** Sample descriptives.

|  | Mean (SD) / Proportion (%) |  | Total |
| --- | --- | --- | --- |
|  | Not culturally engaged | Culturally engaged |  |
| Chronological age (years) | 68.0 (9.3) | 64.5 (8.5) | 65.4 (8.8) |
| Subjective age (years) | 58.9 (15.0) | 54.6 (12.1) | 55.7 (13.1) |
| Physiological age (years) | 70.7 (14.4) | 61.2 (14.2) | 63.6 (14.9) |
| Sex |  |  |  |
| Male | 420 (48.3%) | 1,050 (42.3%) | 1,470 (43.9%) |
| Female | 450 (51.7%) | 1,430 (57.7%) | 1,880 (56.1%) |
| Ethnicity |  |  |  |
| White | 858 (98.6%) | 2,444 (98.5%) | 3,302 (98.6%) |
| Minority ethnic groups | 12 (1.4%) | 36 (1.5%) | 48 (1.4%) |
| Working status |  |  |  |
| Not working/retired/unemployed | 663 (76.2%) | 1,465 (59.1%) | 2,128 (63.5%) |
| Working full/part-time | 207 (23.8%) | 1,015 (40.9%) | 1,222 (36.5%) |
| Housing tenure |  |  |  |
| Other | 386 (44.4%) | 809 (32.6%) | 1,195 (35.7%) |
| Outright homeowner | 484 (55.6%) | 1,671 (67.4%) | 2,155 (64.3%) |
| Physically inactive |  |  |  |
| no | 782 (89.9%) | 2,417 (97.5%) | 3,199 (95.5%) |
| yes | 88 (10.1%) | 63 (2.5%) | 151 (4.5%) |
| Alcohol consumption |  |  |  |
| <5 times a week | 655 (75.3%) | 1,826 (73.6%) | 2,481 (74.1%) |
| 5+ times a week | 215 (24.7%) | 654 (26.4%) | 869 (25.9%) |
| Current smoker |  |  |  |
| no | 699 (80.3%) | 2,251 (90.8%) | 2,950 (88.1%) |
| yes | 171 (19.7%) | 229 (9.2%) | 400 (11.9%) |
| Chronic physical condition |  |  |  |
| no | 714 (82.1%) | 2,128 (85.8%) | 2,842 (84.8%) |
| yes | 156 (17.9%) | 352 (14.2%) | 508 (15.2%) |
| Depression (CESD≥3) |  |  |  |
| no | 601 (69.1%) | 2,082 (84.0%) | 2,683 (80.1%) |
| yes | 269 (30.9%) | 398 (16.0%) | 667 (19.9%) |
| Net non-pension wealth (quintiles) |  |  |  |
| 1 - lowest wealth quintile | 296 (34.0%) | 281 (11.3%) | 577 (17.2%) |
| 2 | 224 (25.7%) | 413 (16.7%) | 637 (19.0%) |
| 3 | 157 (18.0%) | 509 (20.5%) | 666 (19.9%) |
| 4 | 131 (15.1%) | 561 (22.6%) | 692 (20.7%) |
| 5 - highest wealth quintile | 62 (7.1%) | 716 (28.9%) | 778 (23.2%) |
| Educational attainment |  |  |  |
| Degree | 23 (2.6%) | 527 (21.2%) | 550 (16.4%) |
| NVQ3 A level/higher education | 171 (19.7%) | 735 (29.6%) | 906 (27.0%) |
| NVQ2/GCE O level or below | 676 (77.7%) | 1,218 (49.1%) | 1,894 (56.5%) |
| No. of ADL limitations |  |  |  |
| 0 | 626 (72.0%) | 2,180 (87.9%) | 2,806 (83.8%) |
| 1 | 123 (14.1%) | 198 (8.0%) | 321 (9.6%) |
| 2+ | 121 (13.9%) | 102 (4.1%) | 223 (6.7%) |
| Self-reported health |  |  |  |
| Poor | 102 (11.7%) | 66 (2.7%) | 168 (5.0%) |
| Fair | 254 (29.2%) | 320 (12.9%) | 574 (17.1%) |
| Good | 281 (32.3%) | 771 (31.1%) | 1,052 (31.4%) |
| Very good | 181 (20.8%) | 871 (35.1%) | 1,052 (31.4%) |
| Excellent | 52 (6.0%) | 452 (18.2%) | 504 (15.0%) |
| BMI |  |  |  |
| BMI<25 | 192 (22.1%) | 741 (29.9%) | 933 (27.9%) |
| BMI 25-30 | 356 (40.9%) | 1,132 (45.6%) | 1,488 (44.4%) |
| BMI>=30 | 322 (37.0%) | 607 (24.5%) | 929 (27.7%) |

### Exposures

For cultural engagement, we used data from three questions asking participants’ frequency of visits to (a) the cinema, (b) an art gallery, exhibition or museum, or (c) the theatre, concerts or opera. For our main outcome index, we considered the “treatment” group to be “attending one or more of the three cultural exposures every few months or more”. We considered the “control” group to be those who were completely unengaged (i.e. “never engaging”). We also tested this independently for each activity. Sensitivity analyses tested an alternative threshold of overall cultural engagement.

### Outcome

A **physiological age acceleration** measure (measured in years) was derived using the principal component analyses (PCA) method, an established method to measure physiological ageing.^29,30^ It was created using clinical indicators pertaining to the cardiovascular system (pulse pressure and systolic blood pressure), respiratory system (forced vital capacity [FVC], forced expiratory volume in one second [FEV]), the inflammatory system (fibrinogen and C-reactive protein [CRP]), the circulatory system (haemoglobin concentration) and the musculoskeletal system (grip strength). All measures were taken at wave 2 in nurse visits (blood-based biomarkers and physiological testing). This index has previously been validated, published, and used in analyses in relation to other exposures.^24^

The full details of physiological age derivation using the PCA method are summarised in Figure 1 and Supplementary Materials (Appendix 1 and Supplementary Figures 1-2 in Appendix 3). To create the physiological age acceleration/deceleration measure, chronological age was subtracted from physiological age; an approach extensively used in previous research.^31–34^ As in previous studies, this means that “accelerated physiological ageing” refers to a chronological age higher than a physiological age rather than necessarily imply a longitudinal change. Negative scores indicate decelerated ageing (younger physiological age relative to chronological age), and positive scores indicate accelerated ageing (older physiological age relative to chronological age). Analyses of this index show its association with incident functional and mobility limitations, memory impairment, and diverse chronic conditions in the ELSA study population indicating its relevance to ageing processes (Supplementary Table 2).

**Figure 1.**
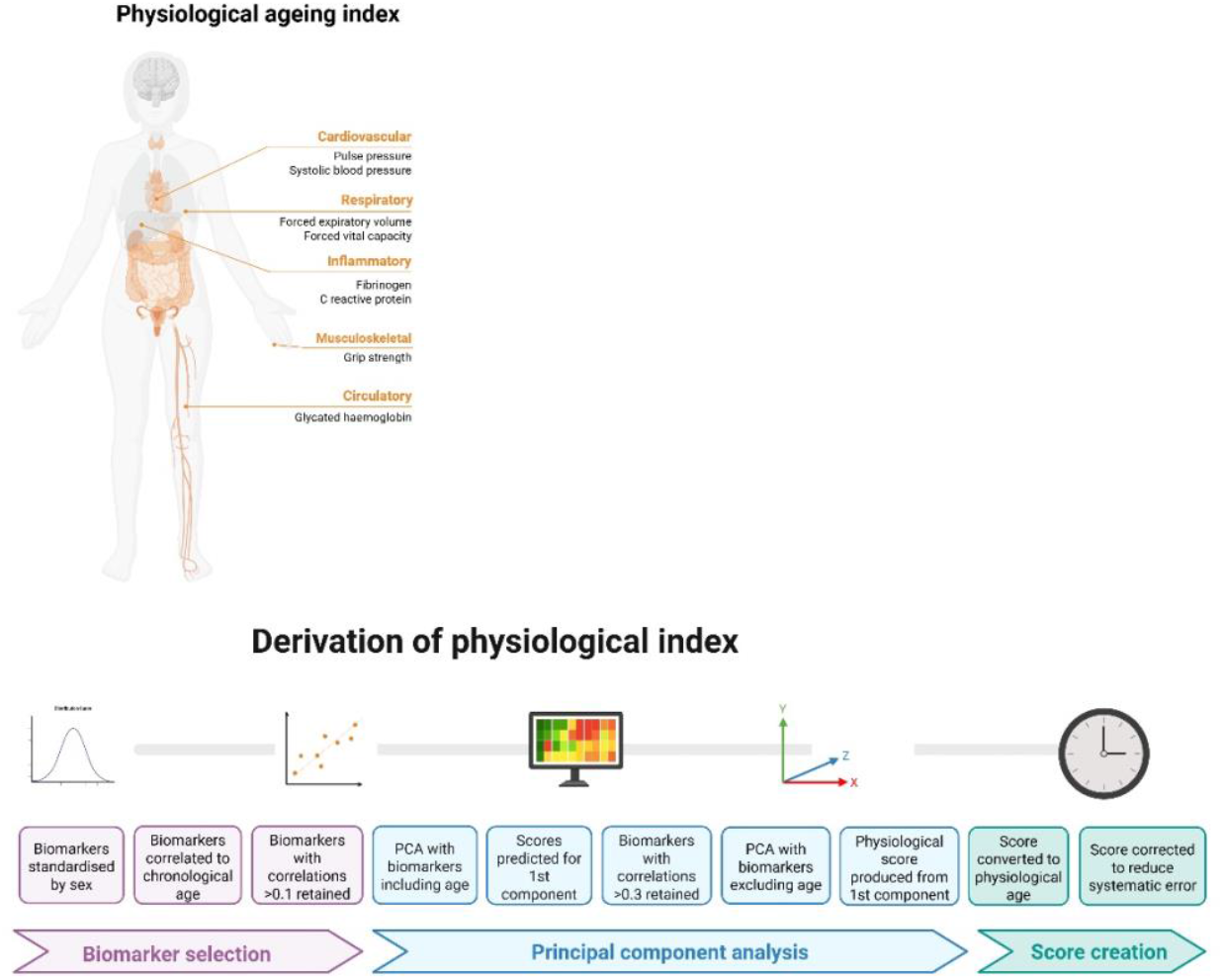
Derivation of physiological ageing index.

As a point of comparison, we also calculated people’s **subjective age** by asking participants how old they felt (measured in years) and subtracting chronological age from this. As a result, negative scores indicate younger subjective age, and positive scores indicate older subjective age. This variable was used to show how physiological age compared both to their chronological age and how old people perceived themselves to be.

### Confounders

Factors identified as predicting both cultural engagement and physiological age were identified using directed acyclic graphs (DAGs) and included as covariates.^35^ Demographic and socio-economic confounders included sex (male vs female), ethnicity (white vs minority ethnic groups), educational attainment (no educational qualifications or qualifications up to age 16 e.g., GCE/O-levels/national vocational qualification [NVQ] 2; qualifications at age 18 [e.g., A-levels]; and higher qualification [e.g., NVQ4/NVQ5/degree]), total non-pension wealth (which combines net financial and physical wealth plus net owner-occupied housing wealth; categorised in quintiles) ^36^, house ownership (whether individuals owned their property outright vs with a mortgage/renting/social housing/other), and employment (not working/retired/unemployed vs working full or part time).

For health and behavioural confounders, we only considered factors that were unlikely to lie on the causal pathway from cultural engagement to ageing, which precluded some behaviours known to be influenced by culture. For example, given cultural engagement provides physical activity through walking incurred in the process of attending events, we only included minimal adjustment for sedentary behaviours (categorised as not engaging in any kind of mild, moderate or vigorous activity in a week). We also measured alcohol consumption (more than 5 days a week vs less), whether participants currently smoked, and whether participants reported currently having a diagnosis of any long-term conditions, including cancer, arthritis, asthma, osteoporosis, Parkinson’s disease, Alzheimer’s disease or dementia, lung disease or cardiovascular disease (including high blood pressure, angina, a previous heart attack, heart failure, a heart murmur, an abnormal rhythm, diabetes, a previous stroke, high cholesterol, or other heart trouble). We additionally looked at the number of difficulties in carrying out activities of daily living that participants reported (ADLs; including dressing, bathing, eating, using a toilet, shopping, taking medications or making telephone calls), self-reported health (poor, fair, good, very good, excellent), BMI (<25, 25-30, ≥30 kg/m^2^), and depression (using the Centre for Epidemiological Studies Depression (CES-D) scale, ≥3). However, as these are behaviours that have much stronger potential roles as partial mediators of effects, we tested the potential influence that including them had within model results in sensitivity analyses (outlined below).

### Statistics

Data were analysed using doubly robust estimation using the inverse-probability-weighted regression adjustment (IPWRA) estimator. This involves building two models to account for the non-random treatment assignment: a regression adjustment model for the outcome and a treatment-assignment model for the exposure, only one of which has to be correctly specified, enhancing the robustness of the analysis.^37^ A strength of this approach is that it provides two opportunities to make valid inferences, instead of just one. IPWRA estimators apply weighted regression coefficients to compute averages of treatment-level predicted outcomes, where the weights are the estimated inverse probabilities of treatment. Following our DAGs, confounders applied to both exposure and outcome were sex, ethnicity, education, wealth, home ownership, employment, physical inactivity, alcohol consumption, smoking, and chronic conditions. Problems with ADLs and self-reported health were additionally applied to the exposure, and BMI and depression to the outcome.

We estimated the average treatment effect in the population (ATE), which is the average difference in outcome if everyone in the population experienced the exposure, versus no-one in the population.^37^ For all analyses, we applied probability weights for the self-completion questionnaire for wave 2 provided in the data, which account for complex sampling strategies and non-response. For RQ1, we looked at overall cultural engagement associations with physiological age cross-sectionally (at wave 2) and then both 4 (wave 4) and 8 (wave 6) years later. As we used doubly robust estimation, we ran these three analyses separately rather than in mixed models. For RQ2, we repeated the cross-sectional analyses for each type of cultural activity separately. Sample size was allowed to vary between analyses to maximise sample.

We also conducted several sensitivity analyses. First, we acknowledge that baseline adjustment of the outcome is controversial as it may lead to downward bias in the presence of residual autocorrelation or overcontrol bias, but equally leaving it out puts analyses at risk of omitted variable bias or overestimation of the effect of the exposure.^38,39^ As such, our core analyses did not control for baseline physiological age, but a sensitivity analysis did adjust for physiological age four years earlier, providing insight into whether cultural engagement relates to *change* in physiological age acceleration over time. Second, depression and BMI could plausibly lie on the causal pathway from cultural engagement to accelerated ageing, so they were only included in the outcome confounder model, not the exposure confounder model, in the main analyses. However, a sensitivity analysis additionally accounted for depression and BMI within the exposure model. A third sensitivity analysis excluded anybody whose physiological age was more than 30 years above or below their chronological age to assess the stability of the findings without the influence of outliers. As doubly-robust estimations do not allow for polytomous exposures (as it works to emulate a trial design with treated and untreated groups), a fourth sensitivity analysis explored if results were stronger when using a more regular frequency of cultural engagement index (never versus monthly instead of every few months or more). A fifth sensitivity analysis dichotomised the results by age (split at 65 years) to ascertain whether results were consistent across ageing. A sixth sensitivity analysis repeated the main analysis using an alternative algorithm to calculate proportional discrepancy in age: age index-chronological age/chronological age.^40^ A final sensitivity analysis considered the stability of results when memory was added into the derivation of the physiological ageing index. Although cognition is not just a physiological process, it was not included in the original index. But cognitive decline provides an additional indicator of ageing from another organ system, extensively researched in accumulating literature brain clocks,^41^ providing a potentially important additional component to considerations of physiological ageing, so we tested to see if it materially affected results.

## Results

### Descriptive statistics

Of the 3,350 adults included in the analyses, the average age was 65.4 years (SD 8.8) and 56.1% were female. Those more engaged in cultural activities had multiple advantages in socioeconomics, health and health behaviours.

Correlations between the different cultural engagement activities included in the model are shown in figure 2a (full correlation matrix for all variables shown in Supplementary Figure 3). Notably, there was no meaningful correlation between physiological age and how old people felt (i.e. their subjective age) (r=0.04) (histogram and heatmap shown in Figures 2c and 2d). Individuals almost exclusively rated their subjective age to be the same or lower than their chronological age (92% the same or below), while physiological age was more evenly distributed above and below chronological age, with a slight skew towards people being older physiologically than chronologically (62% the same or below, figure 2b). There was a slight correlation between older chronological age and lower subjective age and a marked correlation between older chronological age and older physiological age (scatterplots in Figures 2e and 2f; see also Supplementary Figure 4).

**Figure 2.**
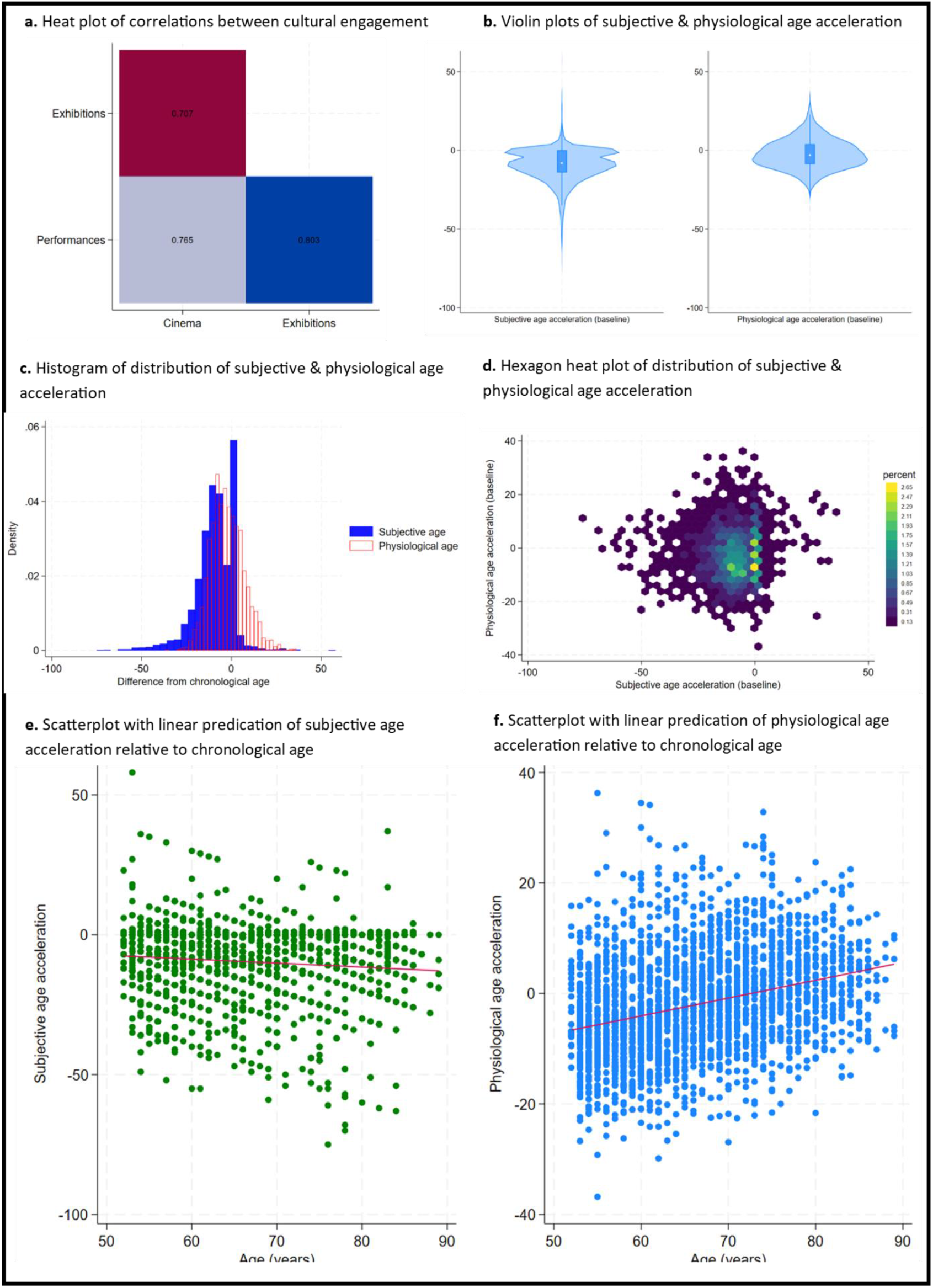
Descriptive figures of exposures and outcomes

**Figure 3.**
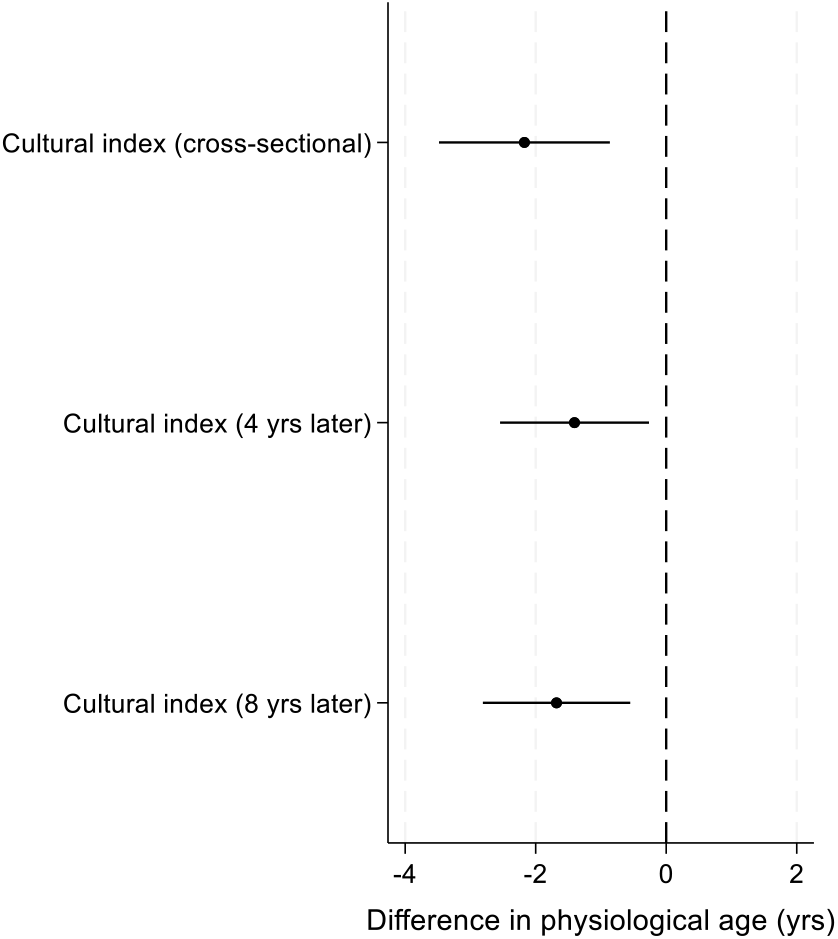
Associations between overall cultural engagement and physiological age acceleration

**Figure 4.**
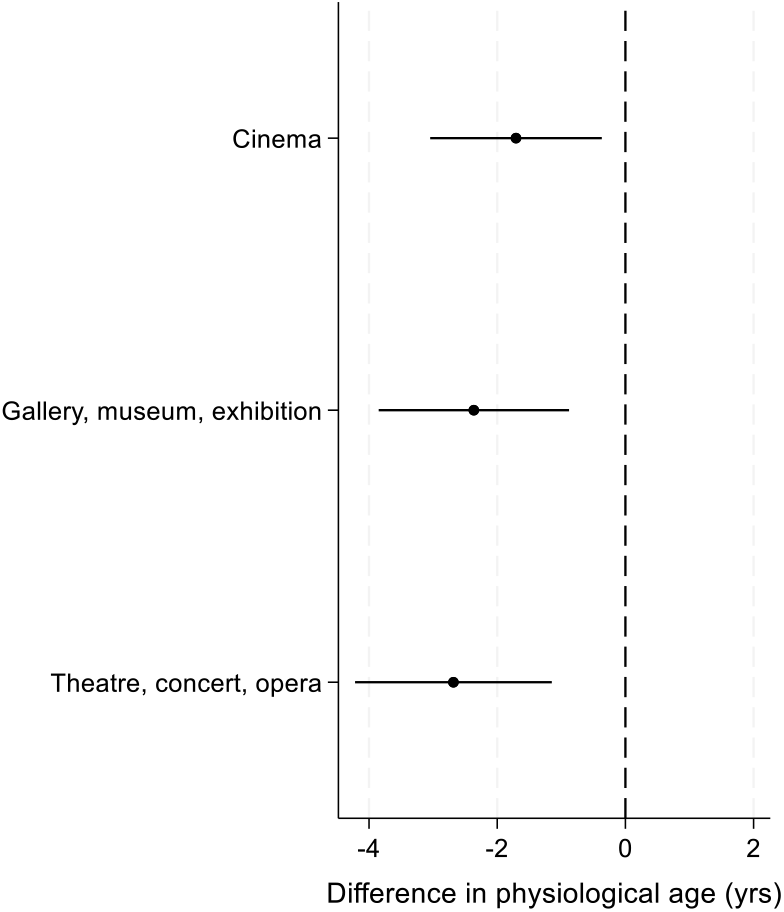
Cross-sectional associations between specific types of cultural activity and physiological age acceleration

### RQ1: overall cultural engagement and physiological age acceleration

Cultural engagement every few months or more was associated with decelerated physiological age (i.e. a lower physiological age relative to chronological age) cross-sectionally, and 4 and 8 years later, even when accounting for confounders within the models (Figure 3; Supplementary Table 4). Those who were regularly engaged in cultural activities had a physiological age 2.2 years younger than those who were unengaged (ATE-2.17; 95% CI-3.48 to-0.86). These effects were maintained 4 and 8 years later (physiological age 1.4 and 1.7 years younger than those who were unengaged respectively), again independent of identified confounders.

### RQ2: specific cultural activities and physiological age acceleration

When considering which type of cultural activity contributed most to the model, all three were associated with slower physiological ageing. Going to performances showed the largest association with decelerated physiological age, with those who went regularly having a physiological age 2.7 years younger than those who never went. Similarly, those who went regularly to the cinema and exhibitions were 2.4 and 1.7 years younger compared with controls respectively.

### Sensitivity analyses

Sensitivity analyses showed that associations at longitudinal follow-up were maintained when controlling for physiological age at baseline, demonstrating that overall cultural engagement was additionally related to a slower *pace* of physiological ageing over the following 4 years of 1.1 years (wave 2-4) and 0.9 years (wave 4-6) (e.g. 4-year follow-up adjusted for baseline: ATE-1.10, 95%CI-2.20 to-0.00, p<.05; Supplementary Figure 5). Adding different confounders into models or excluding outliers also had no material effect (Supplementary Figure 6-7). There was no clear indication of dose response relationships when looking at more frequent engagement (monthly), but sample sizes were smaller in these models, which may have affected the reliability of the specific estimates (Supplementary Figure 8). When considering the analyses stratified by age, associations were strongest for those aged under 65, where overall cultural engagement was related to physiological age deceleration of 2.7 years (95% CI-5.20 to-0.24) compared to 1.2 years (95% CI −2.24 to −0.20) in those aged over 65 (Supplementary Figure 9). When applying an alternative computation of proportional discrepancy in age, findings were materially no different (Supplementary Figure 10). When using continuous variables for plausibly continuous confounders, the results were also comparable (Supplementary Figure 11). Finally, when including memory as part of the physiological ageing index, results were maintained, just slightly stronger (ATE-3.85; 95% CI-5.28 to-2.43; a 37% greater age deceleration than those not engaged; Supplementary Figure 12).

## Discussion

Overall, cultural engagement was related to lower physiological age cross-sectionally and 4 and 8 years later, as well as a decelerated *pace* of ageing over the following four years. The effect was seen consistently for all three types of cultural activity explored (going to the cinema, galleries, museums and exhibitions, and the theatre or live music events). These analyses were robust to multiple sensitivity analyses, including considering alternative confounding structures, outliers, and treatment specification.

The associations between cultural engagement and physiological ageing we found echo previous research that has focused on the individual components of our physiological ageing index, finding reductions in cardiovascular risk factors (e.g., heart rate) and favourable inflammatory profiles.^17,42^ However, our findings extend previous research by demonstrating a relationship between cultural engagement and an aggregate ageing index that combines these phenotypic markers correlated with age and calculates the discrepancy between the resultant physiological age and an individual’s chronological age. As this aggregate index has been independently and longitudinally demonstrated to have predictive potential for diverse pathological age-related health outcomes, we provide the first data indicating that cultural engagement is related to the speed of physiological ageing processes. It is important to note that these findings are likely bidirectional, as for many mechanisms relating cultural engagement with health, with physiological functioning potentially not only influenced by cultural engagement but also affecting one’s ability to engage in cultural activities.^43,44^ However, when we used outcome measures from subsequent years, results were maintained, with only modest attenuation of effect sizes within the first four years of follow-up, and very little further attenuation at eight years of follow-up. Further, the relationship was similarly maintained in longitudinal analyses that additionally adjusted for baseline physiological age, demonstrating that cultural engagement was not only related to decelerated physiological age but also decelerated *pace* of physiological ageing, strengthening causal inference.

Strengths of our study include the use of data drawn from a large, representative sample of older adults, its use of a multi-dimensional index of physiological age that has demonstrated associations with diverse aspects of age-related pathology, and its statistical use of doubly-robust estimators, which provide a clear advance on more traditional regression-based approaches to conditioning on counterfactuals. However, there are several limitations. For example, we were only able to explore cultural engagement, not active participation in the arts, and out measure asked about behaviours in isolation from the contexts in which they occur. As such, the current analyses do not consider whether relationships found are dependent on particular socio-ecological contexts of cultural engagement. Future analyses are recommended that consider broader types of arts engagement and use experimental methods to elucidate how context influences effects, as well as exploring if results differ amongst different demographic subsamples. Additionally, we relied on retrospective reporting of cultural engagement rates over the past year, which could be prone to recall bias, especially amongst older participants.^45^ We used a statistical approach that had many strengths in terms of confounder consideration, but did require considering our exposure as a dichotomous variable to enable the creation of a treatment and non-treatment group. This, naturally, has its own limitations including precluding comprehensive examination of dose-response effects and leading to some attrition (although the sample demographics did not meaningfully change compared to the broader ELSA sample). But it provides an initial step towards future target trial emulation approaches using cultural engagement as an exposure. Additionally, while we included identified confounders and used a statistical method that allows two different model specifications for exposure and outcome (only one of which has to be correct), residual confounding remains possible, so methods such as instrumental variable approaches that better account for unmeasured confounding are also recommended for future work to triangulate with these results and confirm findings. Cross-sectional analyses may be affected by selection bias, and longitudinal analyses attrition bias, if individuals with lower engagement and poorer health are both less likely to survive to participate at baseline and more likely to drop out during follow up. However, this would have attenuated observed associations and therefore does not contradict the findings. Nonetheless, future longitudinal analyses considering survival within models are encouraged. In relation to our physiological ageing index, PCA is a common method for biological age derivation, but it is by no means the only approach and has its own limitations, such as including chronological age within a first component, potentially capturing variance unrelated to ageing, and underestimating the hierarchical nature of biological processes. Nonetheless, we selected PCA above alternatives such as latent variable approaches because even though these other methods better account for hierarchical dependencies, the summary measures produced are not easily interpretable with respect to chronological age. Further, there is a strong precedent for its use in similar derivation of indices.^29,33,46–52^ Future studies are also encouraged to explore how cultural engagement may influence patterns or trajectories of physiological ageing, and whether engaging *consistently* over time is important to effect sizes. An important part of such future work will be able to explore whether temporal variation in confounders affects outcomes, whether effects are bidirectional (as we hypothesise they likely are), and whether feedback effects are evident. Additionally for future work, while this study focused on an overall index of physiological age, in keeping with other ageing research,^29,33,46–52^ studies are also encouraged to look at organ-specific ageing to ascertain whether cultural engagement affects different biological systems at different rates.

Overall, these findings are important as they provide insight into how cultural engagement may be related to physiological processes of ageing. From an intervention design perspective, monthly cultural engagement compared to every few months did not lead to stronger outcomes, suggesting that there may a minimum amount of cultural engagement necessary, beyond which further engagement does not yield stronger benefits. Further, the strength of the relationship with physiological ageing appears stronger amongst those aged 50-65. These details could help to guide the design and implementation of future experimental studies.

## Supporting information

Fancourt_2025_Supplement

## Data Availability

Data is available via the UKDS: https://datacatalogue.ukdataservice.ac.uk/series/series/200011?id=200011#abstract

## Acknowledgements

This research was supported by a Wellcome Trust Discovery Award [326117/Z/25/Z], UK Research and Innovation [MR/Y01068X/1] and the EpiArts Lab-a National Endowment for the Arts Research Lab at the University of Florida, supported in part by an award from the National Endowment for the Arts (1936473-38-24). The opinions expressed are those of the authors and do not represent the views of the National Endowment for the Arts Office of Research & Analysis or the National Endowment for the Arts. The National Endowment for the Arts does not guarantee the accuracy or completeness of the information included in this material and is not responsible for any consequences of its use. The EpiArts Lab is also supported by Americans for the Arts, Bloomberg Philanthropies (F024567), the Dharma Endowment Foundation, the Pabst Steinmetz Foundation, and the State of Florida Division of Arts and Culture (24.c.ne.900.834).

## References

1. Fancourt, D. & Finn, S. What Is the Evidence on the Role of the Arts in Improving Health and Well-Being? A Scoping Review. (2019).

2. Rodriguez, A. K. et al. Arts Engagement as a Health Behavior: An Opportunity to Address Mental Health Inequities. Community Health Equity Res Policy 2752535X231175072 (2023) doi:10.1177/2752535X231175072.

3. Sonke, J. et al. Defining “Arts Participation” for Public Health Research. Health Promotion Practice 15248399231183388 (2023) doi:10.1177/15248399231183388.

4. Warran, K., Burton, A. & Fancourt, D. What are the active ingredients of ‘arts in health’ activities? Development of the INgredients iN ArTs in hEalth (INNATE) Framework. Wellcome Open Res 7, 10 (2022).

5. Fancourt, D., Aughterson, H., Finn, S., Walker, E. & Steptoe, A. How leisure activities affect health: a narrative review and multi-level theoretical framework of mechanisms of action. The Lancet Psychiatry 8, 329–339 (2021).

6. Fancourt, D. & Steptoe, A. Comparison of physical and social risk-reducing factors for the development of disability in older adults: a population-based cohort study. J Epidemiol Community Health 73, 906–912 (2019).

7. Fancourt, D. & Steptoe, A. Physical and Psychosocial Factors in the Prevention of Chronic Pain in Older Age. J Pain 19, 1385–1391 (2018).

8. Fancourt, D. & Tymoszuk, U. Cultural engagement and incident depression in older adults: evidence from the English Longitudinal Study of Ageing. Br J Psychiatry 214, 225–229 (2019).

9. Rogers, N. T. & Fancourt, D. Cultural Engagement Is a Risk-Reducing Factor for Frailty Incidence and Progression. J Gerontol B Psychol Sci Soc Sci 75, 571–576 (2020).

10. Wang, S., Mak, H. W. & Fancourt, D. Arts, mental distress, mental health functioning & life satisfaction: fixed-effects analyses of a nationally-representative panel study. BMC Public Health 20, 208 (2020).

11. Johansson, S.-E., Jansåker, F., Sundquist, K. & Bygren, L. O. A longitudinal study of the association between attending cultural events and coronary heart disease. Communications medicine 3, 72 (2023).

12. Wang, X. et al. Art Engagement and Risk of Type 2 Diabetes: Evidence From the English Longitudinal Study of Ageing. Int J Public Health 68, 1605556 (2023).

13. Bygren, L. O., Konlaan, B. B. & Johansson, S. E. Attendance at cultural events, reading books or periodicals, and making music or singing in a choir as determinants for survival: Swedish interview survey of living conditions. BMJ 313, 1577–1580 (1996).

14. Jensen, A., Pirouzifard, M. & Lindström, M. Arts and culture engagement and mortality: A population-based prospective cohort study. Scand J Public Health 52, 511–520 (2024).

15. Fancourt, D., Steptoe, A. & Cadar, D. Cultural engagement and cognitive reserve: museum attendance and dementia incidence over a 10-year period. The British Journal of Psychiatry 1–3 (2018).

16. Fancourt, D., Steptoe, A. & Cadar, D. Community engagement and dementia risk: time-to-event analyses from a national cohort study. J Epidemiol Community Health 74, 71–77 (2020).

17. Fancourt, D. & Steptoe, A. Cultural engagement predicts changes in cognitive function in older adults over a 10 year period: findings from the English Longitudinal Study of Ageing. Sci Rep 8, 10226 (2018).

18. Fancourt, D. Advancing observational research on arts and health: theory-informed approaches using the RADIANCE framework. Preprint at 10.31234/osf.io/s826r (2024).

19. de Witte, M. et al. The Effects of Arts-Based Interventions in the Treatment and Management of Non-Communicable Diseases: An Umbrella Review and Meta-Analyses. https://europepmc.org/article/ppr/ppr983719 (2025).

20. Moaddel, R. et al. Proteomics in aging research: A roadmap to clinical, translational research. Aging Cell 20, e13325 (2021).

21. Crimmins, E. M., Thyagarajan, B., Kim, J. K., Weir, D. & Faul, J. Quest for a summary measure of biological age: the health and retirement study. GeroScience 43, 395–408 (2021).

22. Tian, Y. E. et al. Heterogeneous aging across multiple organ systems and prediction of chronic disease and mortality. Nat Med 29, 1221–1231 (2023).

23. Crimmins, E. M., Thyagarajan, B., Kim, J. K., Weir, D. & Faul, J. Quest for a summary measure of biological age: the health and retirement study. Geroscience 43, 395–408 (2021).

24. Bloomberg, M. & Steptoe, A. Sex and education differences in trajectories of physiological ageing: longitudinal analysis of a prospective English cohort study. 2025.01.06.25320036 Preprint at 10.1101/2025.01.06.25320036 (2025).

25. Fancourt, D., Bloomberg, M. & Steptoe, A. Social connections are differentially related to perceived and physiological age acceleration amongst older adults. medRxiv 2025–02 (2025).

26. Steptoe, A., Breeze, E., Banks, J. & Nazroo, J. Cohort Profile: The English Longitudinal Study of Ageing. Int J Epidemiol 42, 1640–1648 (2013).

27. Galea, S. & Hernán, M. A. Win-Win: Reconciling Social Epidemiology and Causal Inference. American Journal of Epidemiology 189, 167–170 (2020).

28. Rubin, D. B. Causal Inference Using Potential Outcomes. Journal of the American Statistical Association 100, 322–331 (2005).

29. Nakamura, E., Miyao, K. & Ozeki, T. Assessment of biological age by principal component analysis. Mechanisms of ageing and development 46, 1–18 (1988).

30. Jia, L., Zhang, W. & Chen, X. Common methods of biological age estimation. CIA Volume 12, 759–772 (2017).

31. Liu, N. et al. Association between cardiometabolic index and biological ageing among adults: a population-based study. BMC Public Health 25, 879 (2025).

32. Li, X. et al. Accelerated aging mediates the associations of unhealthy lifestyles with cardiovascular disease, cancer, and mortality. J American Geriatrics Society 72, 181–193 (2024).

33. Dalecka, A., Bartoskova Polcrova, A., Pikhart, H., Bobak, M. & Ksinan, A. J. Living in poverty and accelerated biological aging: evidence from population-representative sample of U.S. adults. BMC Public Health 24, 458 (2024).

34. Husted, K. L. S. et al. A Model for Estimating Biological Age From Physiological Biomarkers of Healthy Aging: Cross-sectional Study. JMIR Aging 5, e35696 (2022).

35. Sauer, B. & VanderWeele, T. J. Use of directed acyclic graphs. in Developing a protocol for observational comparative effectiveness research: a user’s guide (Agency for Healthcare Research and Quality (US), 2013).

36. Banks, J. et al. Financial circumstances, health and well-being of the older population in England: ELSA 2008 (Wave 4). https://www.ifs.org.uk/publications/5315 (2010).

37. Funk, M. J. et al. Doubly Robust Estimation of Causal Effects. American Journal of Epidemiology 173, 761– 767 (2011).

38. Keele, L. & Kelly, N. J. Dynamic models for dynamic theories: The ins and outs of lagged dependent variables. Political analysis 14, 186–205 (2006).

39. Hammerton, G. & Munafò, M. R. Causal inference with observational data: the need for triangulation of evidence. Psychol Med 51, 563–578 (2021).

40. Stephan, Y., Sutin, A. R. & Terracciano, A. How old do you feel? The role of age discrimination and biological aging in subjective age. PLoS One 10, e0119293 (2015).

41. Moguilner, S. et al. Brain clocks capture diversity and disparities in aging and dementia across geographically diverse populations. Nat Med 30, 3646–3657 (2024).

42. Walker, E. S., Fancourt, D., Kumari, M. & McMunn, A. Cross-sectional associations between patterns of cultural engagement and indicators of biological dysregulation. Annals of Human Biology 51, 2399276 (2024).

43. Mak, H. W., Hu, Y., Bu, F., Bone, J. K. & Fancourt, D. Art for health’s sake or health for art’s sake: Disentangling the bidirectional relationships between arts engagement and mental health. PNAS Nexus 3, pgae465 (2024).

44. Fancourt, D. & Bone, J. Advancing observational research on arts and health: theory-informed approaches using the RADIANCE framework. OSF https://osf.io/preprints/psyarxiv/s826r (2024).

45. Rhodes, S., Greene, N. R. & Naveh-Benjamin, M. Age-related differences in recall and recognition: a meta-analysis. Psychon Bull Rev 26, 1529–1547 (2019).

46. Nakamura, E. & Miyao, K. Further evaluation of the basic nature of the human biological aging process based on a factor analysis of age-related physiological variables. J Gerontol A Biol Sci Med Sci 58, 196–204 (2003).

47. Bai, X. et al. Evaluation of biological aging process - a population-based study of healthy people in China. Gerontology 56, 129–140 (2010).

48. Jee, H. et al. Development and application of biological age prediction models with physical fitness and physiological components in Korean adults. Gerontology 58, 344–353 (2012).

49. Zhang, W.-G. et al. Select aging biomarkers based on telomere length and chronological age to build a biological age equation. Age (Dordr) 36, 9639 (2014).

50. Park, J., Cho, B., Kwon, H. & Lee, C. Developing a biological age assessment equation using principal component analysis and clinical biomarkers of aging in Korean men. Arch Gerontol Geriatr 49, 7–12 (2009).

51. Nakamura, E. & Miyao, K. Sex differences in human biological aging. J Gerontol A Biol Sci Med Sci 63, 936– 944 (2008).

52. Nakamura, E., Moritani, T. & Kanetaka, A. Effects of habitual physical exercise on physiological age in men aged 20-85 years as estimated using principal component analysis. Eur J Appl Physiol Occup Physiol 73, 410–418 (1996).

