## Supplementary material for "Cultural engagement is related to decelerated physiological age: doubly-robust estimations in a national cohort study": Fancourt_2025_Supplement

### Appendix 1: Supplemental methods

#### Appendix 1a. Procedures for biomarker collection and inclusion criteria

Methods for blood-based biomarker processing are in line with standard procedures and described in detail elsewhere.1 Participants with CRP concentrations >20 mg/l were excluded as this can indicate acute infection.2 Reproduced from ELSA nurse visit documentation. Available in full at https://www.elsa-project.ac.uk/study-documentation.

**Blood pressure**- All respondents were eligible to have their blood pressure measured. Three measurements were taken of systolic and diastolic pressure as well as pulse rate on the respondent's right arm while they were seated. The mean value of the valid measurements was included in the analyses. Readings were considered invalid if the participant had eaten, drank, smoked, or exercised in the last 30 minutes.

**Grip strength**- All respondents were eligible to have their grip strength measured. Grip strength was evaluated using a Smedley dynamometer. Three measures of grip strength in each hand were taken, with the maximum value in the dominant hand retained for analysis.

**Waist circumference**- All respondents were eligible to have their waist measurements taken, unless they were chair-bound or had a colostomy or ileostomy. Measurements were taken twice; however, if the second measurement differed from the first by 3cm or more, the nurse received an error message in the CAPI program and was prompted to either amend one of the previous responses if a mistake had been made entering a measurement, or to take a third measurement. If the nurse believed that the measurements they took were 0.5cm more or less than the true measurement because of problems encountered (e.g. clothing the respondent was wearing), this was considered unreliable.

**Blood samples**- All sample members who gave consent were eligible for a blood sample to be taken. The only exceptions to this were people with clotting or bleeding disorders, people with a history of fits or convulsions, or people who were on anticoagulant drugs. Respondents aged 80 or under were asked to fast before their nurse visit so a fasting blood sample could be taken. Respondents were not asked to fast if they had diabetes and were on treatment or if they were considered to be malnourished or otherwise unfit to fast (this information was obtained from the interviewer). Respondents who were asked to fast were given guidelines about when and what they could eat based on their appointment time. In the nurse visit, respondents were asked when they had last eaten and, if this was in the last 24 hours, what they had eaten. The CAPI program used their responses to work out if they had fasted adequately. A respondent was considered to have fasted and therefore be eligible for a fasting blood sample if they hadn't eaten or drunk anything (apart from water) on the day of their nurse visit OR they hadn't eaten or drunk anything (apart from water) in the past 5 hours and had only had a light meal (see appointment record card) or a piece of fruit or drink the last time they ate.

**Lung function**- Three measurements each were taken of forced vital capacity and forced expiratory volume using a NDD Easy On-PC spirometer, with the largest measure retained for analysis. All respondents were eligible to have their lung function measured, except for the following: (i) those who had had abdominal or chest surgery in the preceding 3 weeks (ii) those who had been admitted to hospital with a heart complaint in the preceding 6 weeks (iii) Those who had had eye surgery in the preceding 4 weeks.

#### Appendix 1b. Derivation of physiological ageing index

Physiological age was derived based on the principal component analysis (PCA) method, which is among the most commonly used methods of biological age derivation.[1–3] The concept underlying this method is that the ageing of multiple organ systems can be represented as a linear combination of clinical markers that capture changes in organ function with age. By selecting clinical markers representing different organ systems that are associated with chronological age in a healthy subsample of the study population, PCA can be used to produce a score that represents the healthy ageing process of these organ systems. The weights derived from the healthy subpopulation are then used to produce physiological age in the wider study population.

Derivation of physiological age was performed using the original ELSA cohort respondents only without including refreshment cohorts (N=8,253). We first excluded 7,182 participants reporting diabetes, cancer, lung disease, cardiovascular conditions, stroke, arthritis, asthma, high blood pressure, high cholesterol, Parkinson's disease, osteoporosis, or dementia, and 249 participants who were missing biomarker data, leaving 822 healthy participants in the derivation sample.

We standardised each biomarker by sex and examined correlations between the biomarker and chronological age at each wave, retaining those with a correlation coefficient of ≥0.10 at any wave. This led us to retain pulse, systolic blood pressure, fibrinogen, C-reactive protein, glycated haemoglobin, forced expiratory volume (FEV), forced vital capacity (FVC), and grip strength for further examination. We then performed PCA including chronological age and the retained biomarkers, predicted scores for the first component, and examined correlations between each biomarker and the predicted score. We retained biomarkers with at least moderate correlation (r≥0.3) with predicted score; we did not exclude any biomarkers at this stage. We performed PCA again with the remaining biomarkers but excluding chronological age. We used the first component to produce a physiological score for all participants, including those initially excluded due to chronic conditions. We converted this physioloogicalscore to physiologicalage using the following formula, which is the standard procedure for derivation of physiological [1]

$$PAi = (PSi * sCA) + \bar{x}CA$$

Where $PAi$ denotes physiological age for individual $i$, $PSi$ denotes the physiological score for individual $i$, $sCA$ denotes the derivation sample standard deviation of chronological age and $\bar{x}CA$ denotes the derivation sample mean chronological age. We applied the following correction to physiological age to reduce systematic error:[3,4]

$$PAi = PAi + [(CAi - \bar{x}CA) \times (1 - b)]$$

Where $CAi$ denotes chronological age for individual $i$, and $b$ denotes the simple linear regression coefficient of $PA$ on $CA$ in the derivation sample.

#### Appendix 1c. Outcome ascertainment and Cox proportional hazards models for internal validation

Participants were asked to report functional limitations lasting at least three months. ADL included walking across a room, dressing, bathing, eating, getting in/out of bed, and toileting. IADL included using a map, using the telephone, managing money, taking medications, grocery shopping, preparing a hot meal, and doing work around the house/garden. Mobility activities included walking 100 yards, sitting for two hours, getting up from a chair, climbing one flight of stairs, stooping/kneeling/crouching, lifting/carrying 10 lbs, picking up a coin, reaching/extending the arms, or pushing/pulling a large object. For a given wave, a participant was considered to have ADL limitations if they reported at least "some difficulty" with one or more ADL, IADL limitations if they reported "some difficulty" with one or more IADL, and mobility limitations if they reported "some difficulty" with one or more mobility activities.

To assess memory (which was used in sensitivity analyses only), participants were administered Consortium to Establish a Registry for Alzheimer's Disease immediate and delayed recall tasks,[7] in which participants are read a 10-word list and asked to recall it immediately and after a delay. Immediate and delayed recall scores are summed. For a given wave, participants were considered to show evidence of memory impairment if their memory score was >1.5 standard deviations below the mean for their five-year age group. This cut-off is widely used to identify cognitive impairment when clinical evaluation is not possible.[8]

At each interview participants were also asked to report whether they'd received a diagnosis of diabetes, lung disease, cardiovascular disease, stroke, high cholesterol, high blood pressure, cancer, arthritis, osteoporosis, dementia, or Parkinson's disease.

Data from waves 2-10 were used to validate physiological ageing. Physiological age derived from the first biomarker measurement for each participant was used to produce ageing acceleration (physiological age – chronological age). We then used Cox proportional hazards models to examine associations between ageing acceleration and incident ageing-related health outcomes. These models had a time scale in years since measurement of ageing acceleration. The maximum follow-up period comprised nine waves or 19 years of follow-up (2004/05-2021/23). Separate models were fitted for each outcome. Multiple-event-per-subject models were used for functional limitations and memory impairment; single-event-per-subject models were used for chronic conditions. Data were censored at the interview of record of the outcome (single-event-per-subject only) or the end of follow-up (December 2021/23). Models were adjusted for chronological age and sex with the proportional hazards assumption evaluated using Schoenfeld residuals.

### Appendix 2: Supplemental tables

#### Supplementary Table 1: Sample size across waves

| | N |
|---|---|
| Physiological age acceleration (baseline) | 2,607 |
| Physiological age acceleration (4 year f-up) | 1,840 |
| Physiological age acceleration (8 year f-up) | 1,477 |

#### Supplementary Table 2. Associations between physiological age acceleration and ageing-related outcomes

| | HR for 5-year change in ageing acceleration (95% CI) | *P-value* |
|---|---|---|
| *Functional limitations* | | |
| ADL | 1.15 (1.14-1.17) | <0.0001 |
| IADL | 1.16 (1.15-1.17) | <0.0001 |
| Mobility activities | 1.09 (1.09-1.10) | <0.0001 |
| Memory impairment | 1.16 (1.14-1.17) | <0.0001 |
| *Chronic conditions* | | |
| Diabetes | 1.34 (1.30-1.38) | <0.0001 |
| Lung disease | 1.23 (1.19-1.28) | <0.0001 |
| Cardiovascular disease | 1.09 (1.06-1.11) | <0.0001 |
| Stroke | 1.16 (1.12-1.21) | <0.0001 |
| High cholesterol | 1.08 (1.06-1.09) | <0.0001 |
| High blood pressure | 1.26 (1.23-1.28) | <0.0001 |
| Cancer | 1.03 (1.00-1.05) | 0.065 |
| Arthritis | 1.06 (1.03-1.08) | <0.0001 |
| Osteoporosis | 1.04 (1.01-1.07) | 0.018 |
| Dementia | 1.09 (1.05-1.14) | <0.0001 |
| Parkinson's disease | 0.98 (0.90-1.07) | 0.630 |

Ageing acceleration corresponds to physiological age – chronological age. Based on Cox proportional hazards models adjusted for sex and chronological age. Outcomes ascertained from waves 2-10 of ELSA (2004/05-2021/23). Abbreviations: HR, hazard ratio; CI, confidence interval; ADL, activities of daily living; IADL, instrumental activities of daily living.

#### Supplementary Table 3. Results of restricted cubic spline tests of nonlinearity for age-biomarker associations.

| Biomarker | P-value* |
|---|---|
| Pulse pressure | 0.032 |
| Systolic blood pressure | 0.71 |
| Fibrinogen | 0.95 |
| C-reactive protein | 0.056 |
| Glycated hemoglobin | 0.16 |
| FEV | 0.42 |
| FVC | <0.0001 |
| Grip strength | <0.0001 |

Models are linear mixed models with a random intercept and slope at the individual level, fitted to biomarker data from waves 2, 4, and 6. P-values are from likelihood ratio tests comparing models with linear versus spline terms for age. Although several tests were statistically significant, spline plots (Supplementary Figure 1) indicated monotonic, approximately linear associations across the analytic age range.

#### Supplementary Table 4. Average treatment effects and 95% confidence intervals for main analyses

| | Physiological age acceleration (ATE + 95% CI; years) |
|---|---|
| **RQ1** | |
| Cross-sectional | -2.17 [-3.48,-0.86] *** |
| 4 years later | -1.41 [-2.55,-0.26] *** |
| 8 years later | -1.68 [-2.81,-0.55] *** |
| **RQ2** | |
| Cinema | -1.71 [-3.05,-0.37] *** |
| Exhibitions | -2.36 [-3.85,-0.88] *** |
| Performances | -2.68 [-4.22,-1.15] *** |

ATE = average treatment effect, CI = confidence interval. * = p<.05, ** p<.=01, *** p<.001. Results reported from fully-adjusted models

### Appendix 3: Supplemental figures

#### Supplementary Figure 1. Restricted cubic spline plots of age–biomarker associations (ages 50-90).

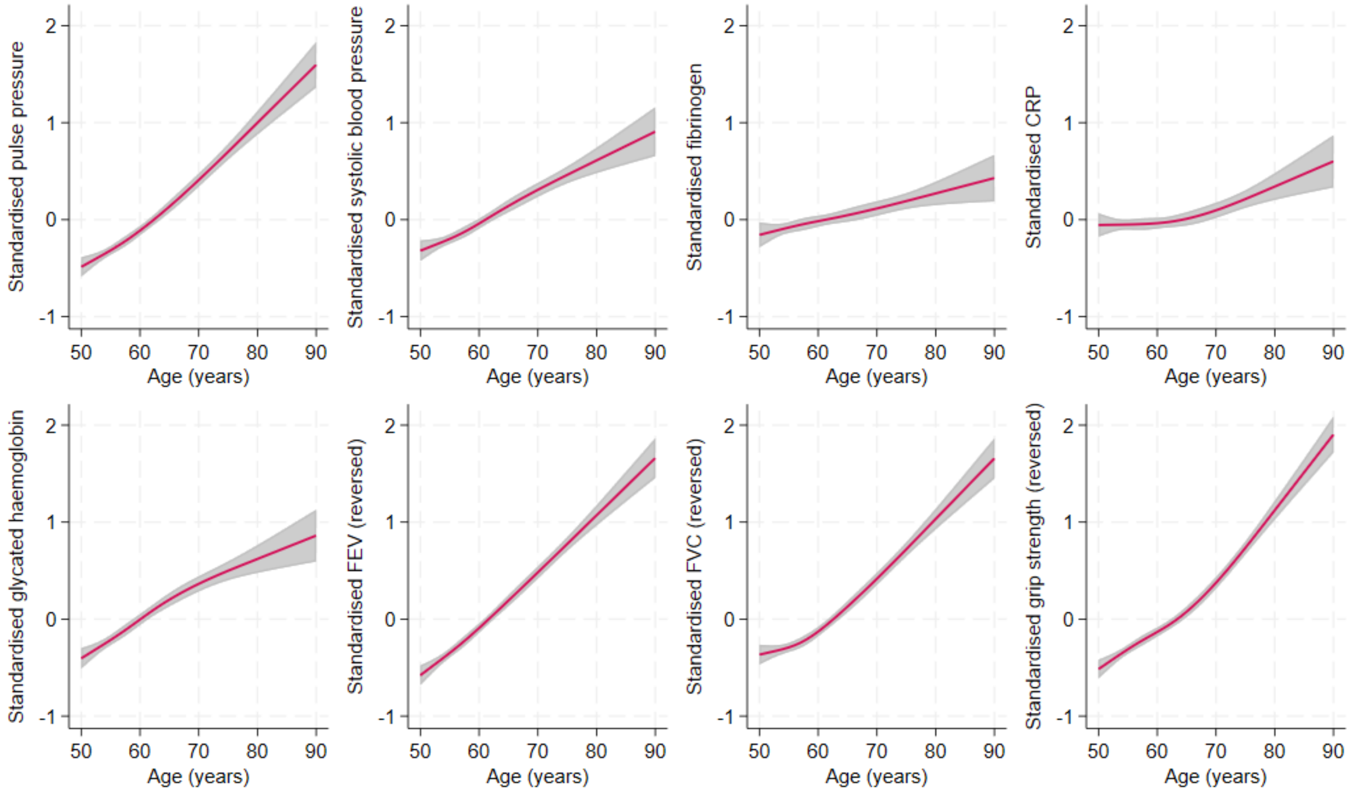

Plots based on linear mixed models with a random intercept and random slope for age at the individual level, fitted to biomarker data from waves 2, 4, and 6. Age was modelled using restricted cubic splines with four knots (at the 5th, 35th, 65th, and 95th percentiles of the analysis sample). Shaded areas represent 95% confidence intervals. Although likelihood ratio tests (Supplementary Table 3) indicated statistically significant departures from linearity for several biomarkers, the spline plots show monotonic, approximately linear associations across the analytic age range.

#### Supplementary Figure 2. Restricted cubic spline plot of the association between chronological age and the PCA-derived subjective age index (ages 50-90).

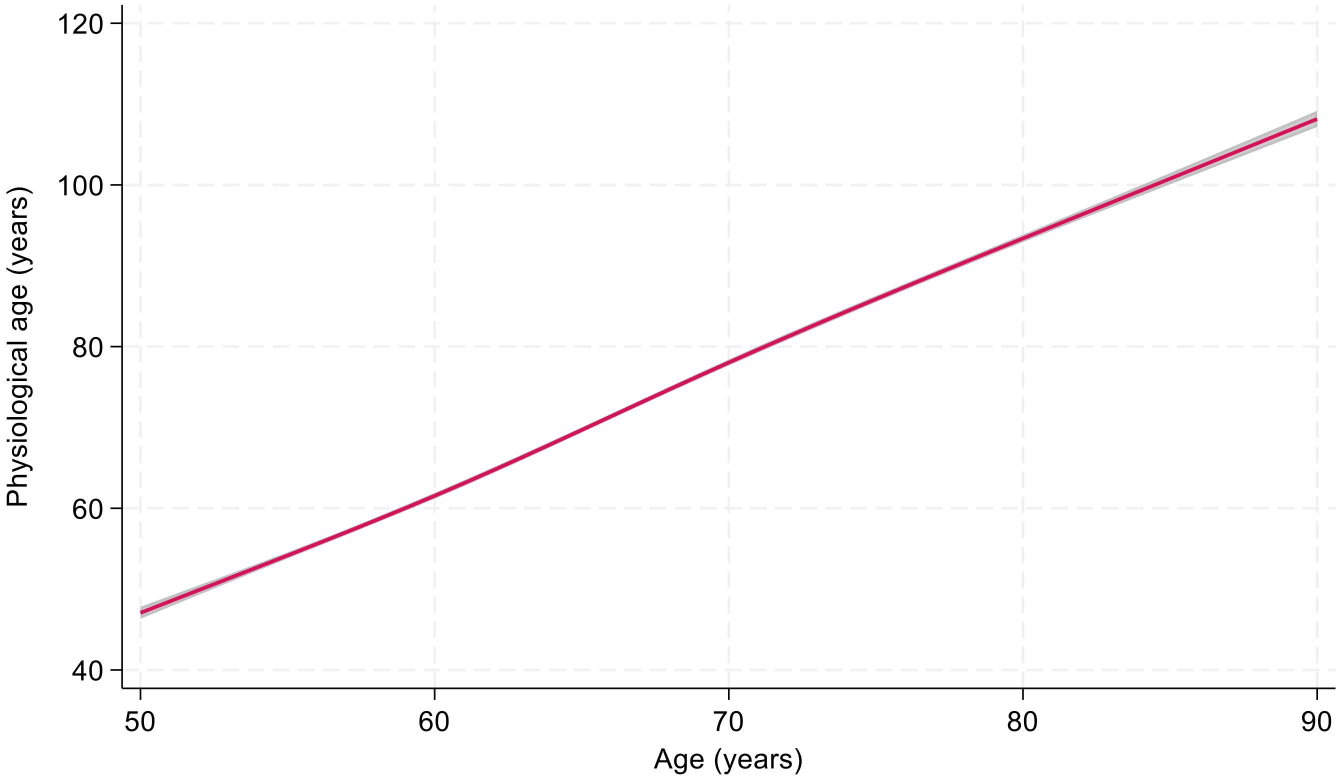

Plot based on linear mixed model with a random intercept and random slope for chronological age at the individual level, fitted to biomarker data from waves 2, 4, and 6. Age was modelled using restricted cubic splines with four knots (at the 5th, 35th, 65th, and 95th percentiles of the analysis sample). Shaded areas represent 95% confidence intervals.

#### Supplementary Figure 3: correlation matrix between all variables in the study

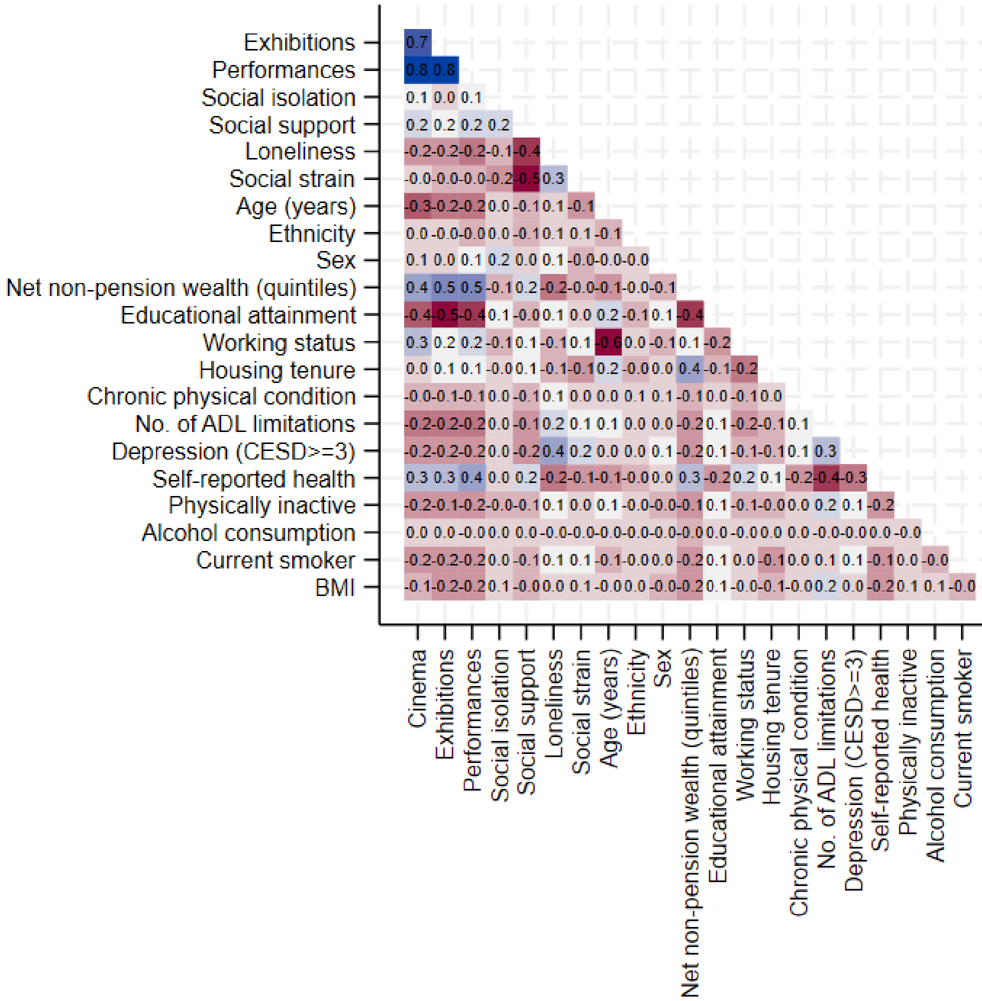

#### Supplementary Figure 4: bivariate histograms using hexagon heat plots to show the relative frequency of associations beween subjective and physiologicalage and chronological age

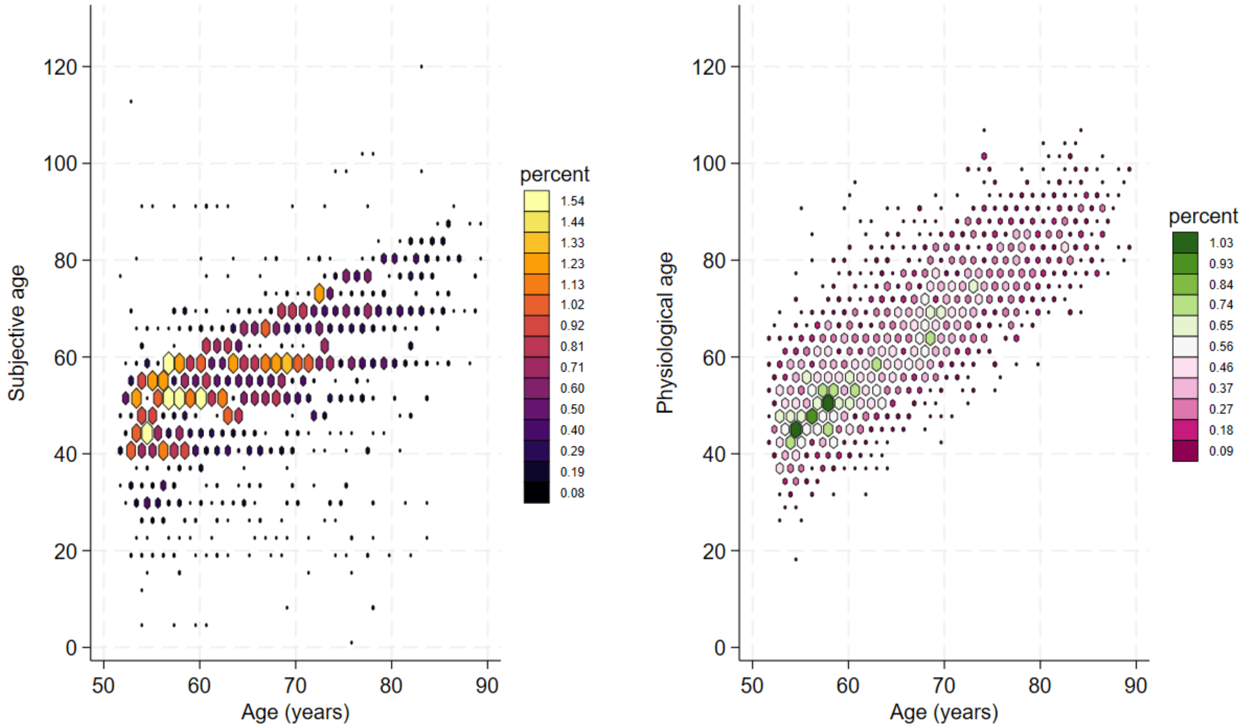

#### Supplementary Figure 5: Associations between overall cultural engagement and perceived age acceleration adjusting for age acceleration four years earlier

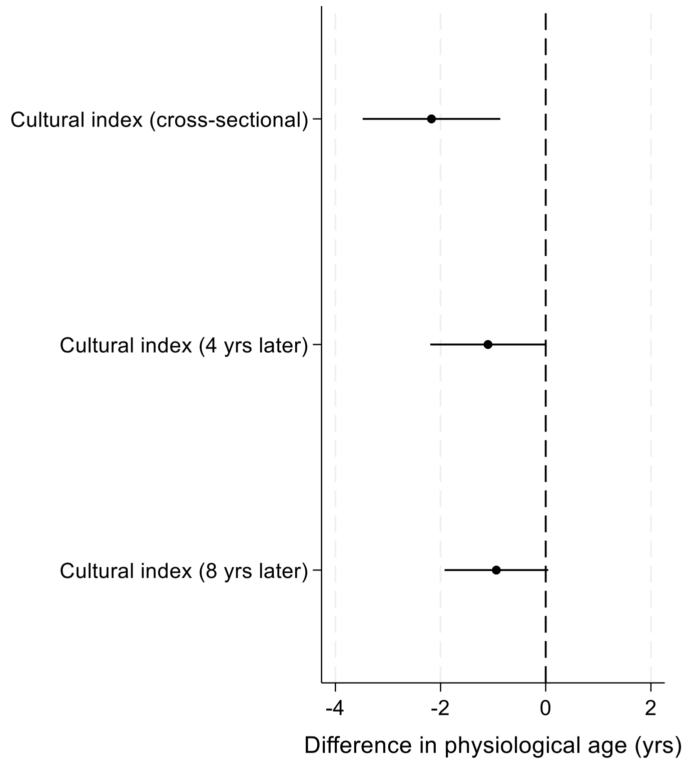

#### Supplementary Figure 6: Associations between overall cultural engagement and physiological acceleration additionally adjusting for BMI and depression in the exposure confounder model

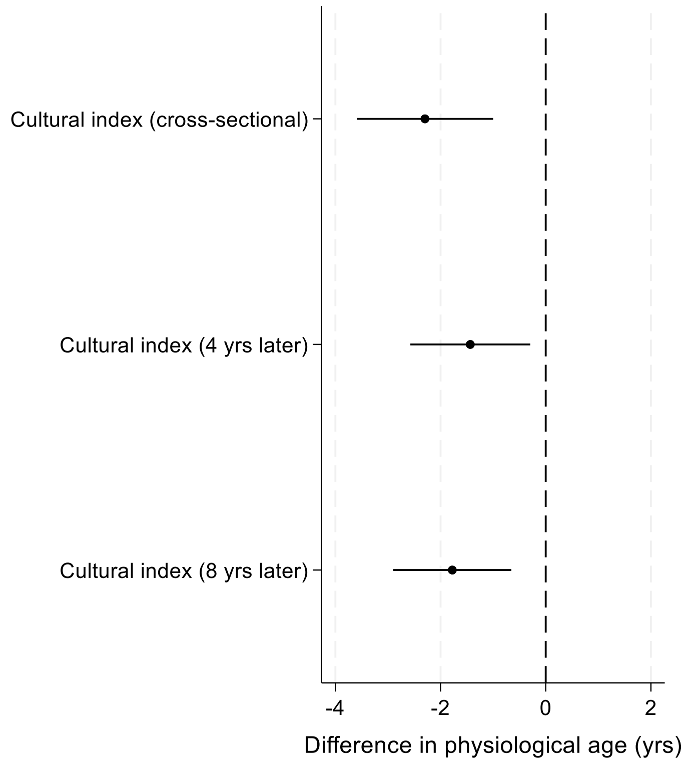

#### Supplementary Figure 7: Associations between overall cultural engagement and physiological age acceleration, removing anybody whose physiological or subjective age was more than 30 years from their chronological age

*Notes: N=2,465; 1,773; 1,449 respectively for cross-sectional, 4 and 8 year follow-up*

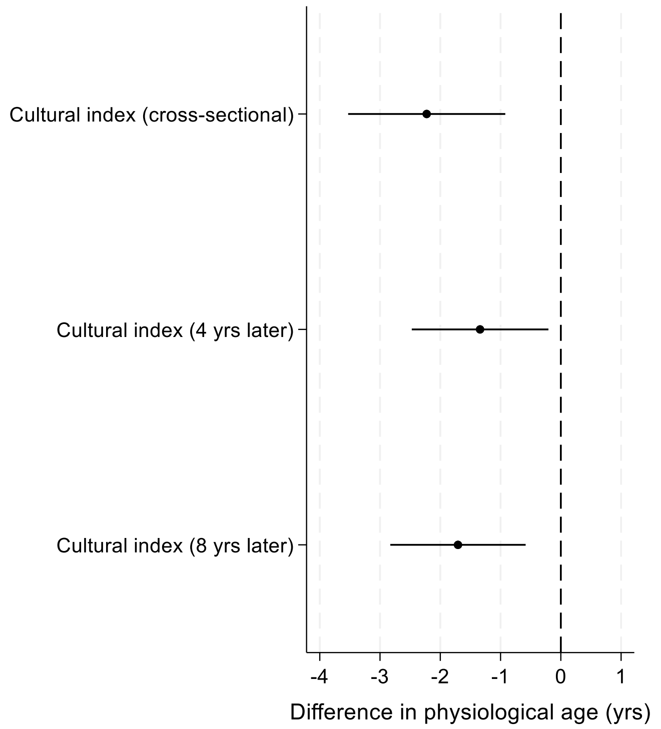

#### Supplementary Figure 8: Associations between overall cultural engagement and physiological age acceleration, using monthly frequency of engagement

*Notes: N=1,656; 1,118; 854 respectively for cross-sectional, 4 and 8 year follow-up*

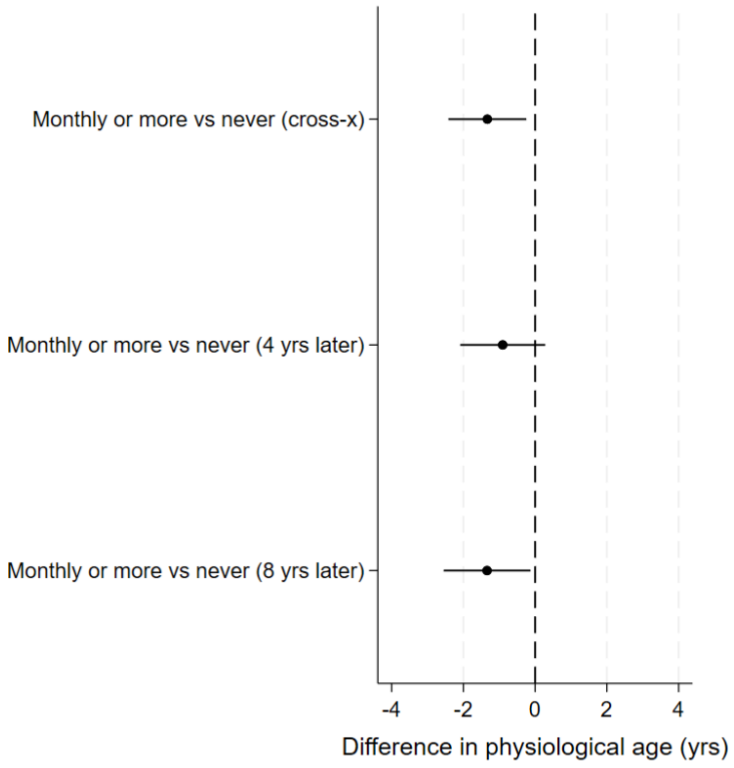

#### Supplementary Figure 9: Associations between overall cultural engagement and physiological age acceleration, split by age

*Notes: Physiological age for adults aged<65, n=1,265; physiological age for adults aged>=65, n=1,330*

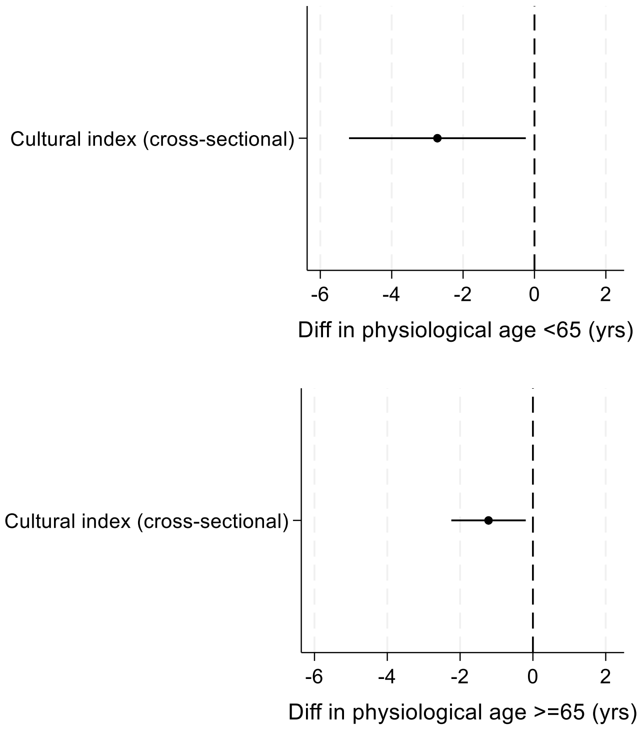

#### Supplementary Figure 10: Associations between overall cultural engagement and physiological age acceleration using an alternative computation of proportional discrepancy in age (age index-chronological age/chronological age).

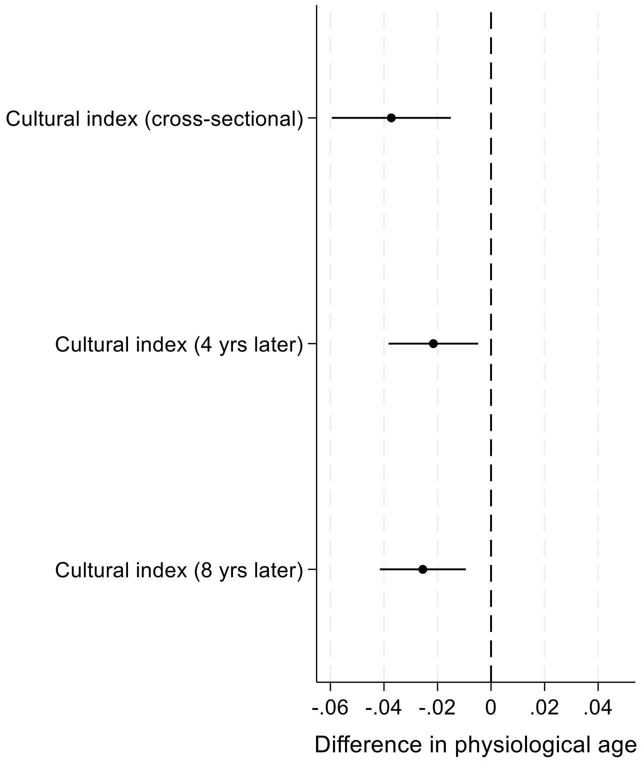

#### Supplementary Figure 11: Associations between overall cultural engagement and physiological age acceleration when including memory too within the physiological ageing index

*Notes: N=2,595; 1,851; 1,486 respectively for cross-sectional, 4 and 8 year follow-up*

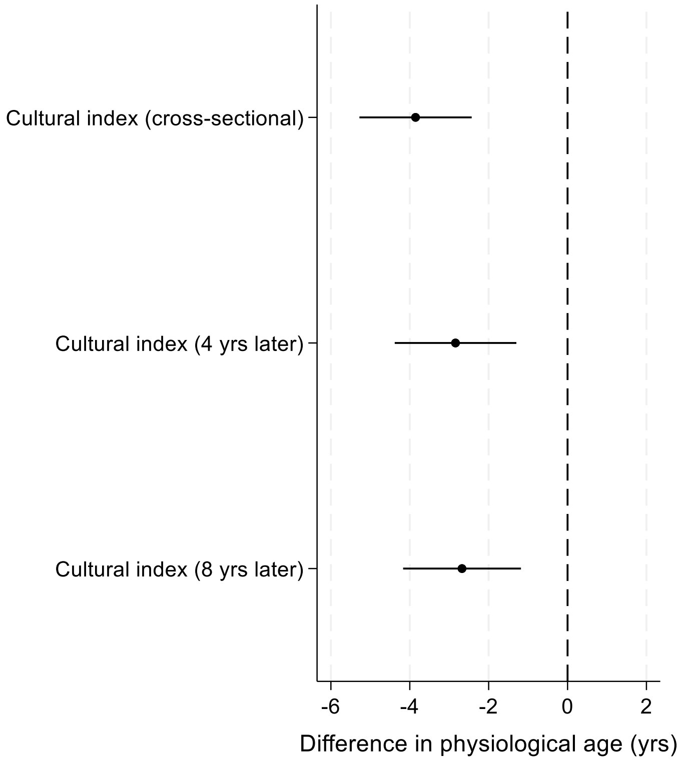

#### Supplemental references

1. Jia, L., Zhang, W. & Chen, X. Common methods of biological age estimation. *CIA* **Volume 12**, 759–772 (2017).
2. Nakamura, E., Miyao, K. & Ozeki, T. Assessment of biological age by principal component analysis. *Mechanisms of ageing and development* **46**, 1–18 (1988).
3. Bafei, S. E. C. & Shen, C. Biomarkers selection and mathematical modeling in biological age estimation. *npj Aging* **9**, 13 (2023).
4. Dubina, T. L., Mints, A. Y. & Zhuk, E. V. Biological age and its estimation. III. Introduction of a correction to the multiple regression model of biological age and assessment of biological age in cross-sectional and longitudinal studies. *Experimental gerontology* **19**, 133–143 (1984).
